# Limit of detection in different matrices of nineteen commercially available rapid antigen tests for the detection of SARS-CoV-2

**DOI:** 10.1101/2021.03.19.21253950

**Authors:** Ana I. Cubas-Atienzar, Konstantina Kontogianni, Thomas Edwards, Dominic Wooding, Kate Buist, Caitlin R. Thompson, Christopher T. Williams, Edward I Patterson, Grant Hughes, Lisa Baldwin, Camille Escadafal, Jilian A. Sacks, Emily R. Adams

## Abstract

In the context of the coronavirus disease 2019 (COVID-19) pandemic there has been an increase of the use of antigen-detection rapid diagnostic tests (Ag-RDT). The performance of Ag-RDT vary greatly between manufacturers and evaluating their analytical limit of detection (LOD) has become high priority. Here we describe a manufacturer-independent evaluation of the LOD of 19 marketed Ag-RDT using live SARS-CoV-2 spiked in different matrices: direct culture supernatant, a dry swab, and a swab in Amies. Additionally, the LOD using dry swab was investigated after 7 days’ storage at −80°C of the SARS-CoV-2 serial dilutions. An LOD of ≈ 5.0 × 10^2^ pfu/ml (1.0 × 10^6^ genome copies/ml) in culture media is defined as acceptable by the World Health Organization. Fourteen of nineteen Ag-RDTs (ActiveXpress, Espline, Excalibur, Innova, Joysbio, Mologic, NowCheck, Orient, PanBio, RespiStrip, Roche, Standard-F, Standard-Q and Sure-Status) exceeded this performance criteria using direct culture supernatant applied to the Ag-RDT. Six Ag-RDT were not compatible with Amies media and a decreased sensitivity of 2 to 20-fold was observed for eleven tests on the stored dilutions at −80°C for 7 days. Here, we provide analytical sensitivity data to guide appropriate test and sample type selection for use and for future Ag-RDT evaluations.

## Introduction

During the Severe Acute Respiratory Syndrome Coronavirus 2 (SARS-CoV-2) pandemic, reverse transcription polymerase chain reaction (RT-PCR) has become the gold standard for diagnosis of acute infection^1^. However, RT-PCR technologies have several limitations: they are not deployed easily, require significant laboratory infrastructure, reagents, and skilled staff, and during this pandemic shortages in global supply have presented challenges^2–4^. In addition, the turnaround time from sample collection to result can be up to 72 hours compromising the effectiveness of triage, isolation, and contact tracing strategies^5,6^. In comparison, rapid diagnostic tests (RDTs) based on antigen detection (Ag-RDT) can determine the presence of the virus in a clinical sample on site in less than 30 minutes without the need for a laboratory. Ag-RDTs are faster, cheaper and can be available at the point-of-care (POC), which is especially important for implementation in community and low-resource settings, where limited laboratories and trained staff are available, and there may be suboptimal cold chain capacity to ensure appropriate conditions for more complex testing^7–9^. Furthermore, their use could also enable rapid isolation of cases and their contacts.

Ag-RDTs are less sensitive than RT-PCR, but clinical evaluation data is emerging that demonstrates Ag-RDTs are accurate at detecting the vast majority of individuals with a high-viral load (cycle threshold (Ct) on RT-PCR ≤ 25.0 or >10^6^ genomic virus copies/ml)^7,10–16^. In addition, in outbreak scenarios, a diagnostic test with lower sensitivity but a fast result enables quick interventions such as self-isolation and isolating contacts of cases^17^. Implementation of Ag-RDTs into testing algorithms would allow rapid detection and isolation of new cases and thereby support the test, trace and isolate strategy, aiming to stop transmission chains and reduce the impact of COVID-19.

Ag-RDTs have been recently used for screening asymptomatic people in high prevalence areas and frontline workers to quickly identify persons with a SARS-CoV-2 infection to adapt infection prevention and control measures, thus preventing transmission in the community^18–20^. A mass testing program screening asymptomatic people in Slovakia using Ag-RDTs was shown to reduce the prevalence of SARS-CoV-2 infections by >50% within two weeks^18^.

Despite the increased adoption of Ag-RDTs as an alternative of RT-PCR, independent analytical sensitivity data is currently lacking for many rapid antigen tests. Evaluation of Ag-RDTs using spiked samples in the laboratory before proceeding on clinical specimens is of paramount importance because the sensitivity of Ag-RDTs is highly variable depending on the manufacturer, ranging 0%-95% in respiratory samples^7,21–24^.

Here we describe a single-center, manufacturer-independent analytical validation of nineteen commercially available Ag-RDTs. The aims of the study were to assess the limit of detection (LOD) using viral culture in different sample matrices: direct culture supernatant, dry swab and swabs in Amies. The effect on the LOD of one freeze-thaw cycle following storage at −80°C was also explored; demonstration of adequate performance using this sample type could support future rapid evaluation of Ag-RDTs with stored material.

## Results

### LOD using different matrices

The LOD was evaluated in three matrices: direct culture supernatant, dry swab and swab in Amies. Direct viral culture supernatant was used as it is the standardized protocol for the evaluation of LOD in Ag-RDTs^25^. Dry swab matrix using the proprietary swab kit was selected to evaluate the LOD in the sample type as defined in the instructions for use (IFU). Finally, swab in Amies was used to assess the use of the same swab used for RT-PCR as a sample type for Ag-RDTs.

A predefined performance criterion of an analytical LOD of ≤ 5.0 × 10^2^ plaque forming units (pfu)/ml (≈ 10^6^ genome copy numbers (gcn)/ml) using direct culture supernatant was selected based on current WHO and national standards^25,26^. Fourteen of nineteen Ag-RDTs evaluated in this study had an LOD of ≤ 5.0 × 10^2^ pfu/ml (ActiveXpress, Espline, Excalibur, Innova, Joysbio, Mologic, NowCheck, Orient, PanBio, RespiStrip, Roche, Standard-F, Standard-Q and Sure-Status) using direct culture supernatant and the remaining had an LOD of 1.0-5.0 × 10^3^ pfu/ml (Biocredit, Genedia, iChroma and Wondfo) (Figure 1). Espline, iChroma, Innova Panbio and Roche had the lowest LOD using direct culture supernatant reaching 0.5-1.0 × 10^2^ pfu/ml, followed by ActiveXpress, Excalibur, Mologic, Orient, Standard-Q and Sure-Status and with an LOD of 2.5 × 10^2^ pfu/ml. Biocredit, Genedia, Respi-Strip, Standard-F and Wondfo and were the least sensitive with an LOD of 0.5-5.0 × 10^3^ pfu/ml. See Figure 1 and Supplementary materials for detailed LODs on all Ag-RDTs.

The LOD using dry swab was poorer in all tests when compared to direct culture supernatant with the exception of Mologic, where the LOD was the same using direct culture supernatant and dry swab. The LOD using dry swab was ≤ 5.0 × 10^2^ pfu/ml only in four Ag-RDT (Espline, Mologic, Roche and Sure-Status), 1.0-5.0 × 10^3^ pfu/ml in seven (Bioeasy, Innova, NowCheck, Orient, Panbio, Respi-Strip and Standard-Q) and ≥ 1.0 × 10^4^ pfu/ml in seven (ActiveXpress, Biocredit, Genedia, iChroma, Joysbio, Standard-F and Wondfo). The least sensitive using dry swab matrix was Joysbio with an LOD of 2.5 × 10^5^ pfu/ml (Figure 1).

None of the Ag-RDTs evaluated specifically indicate compatibility with swabs in Amies media. However, four tests recommend the use of universal or viral transport media (UTM/VTM) (Biocredit, Respi-Strip, Roche and Wondfo), three tests do not recommend the use of UTM/VTM (NowCheck, Standard-F and Standard-Q), and the remaining kits do not mention the use of any transport media. Six Ag-RDTs (Excalibur, Joysbio, NowCheck, Orient, Sure-Status and Wondfo) were found to be incompatible with the Amies media, as these showed a positive test line with the negative control sample. Of these, Wondfo is the only kit which recommends the use of transport media. LODs using swabs in Amies media was poorer than using dry swabs except in two test where the LOD was the same as with the dry swabs (PanBio and Standard-Q).

**Figure 1.**
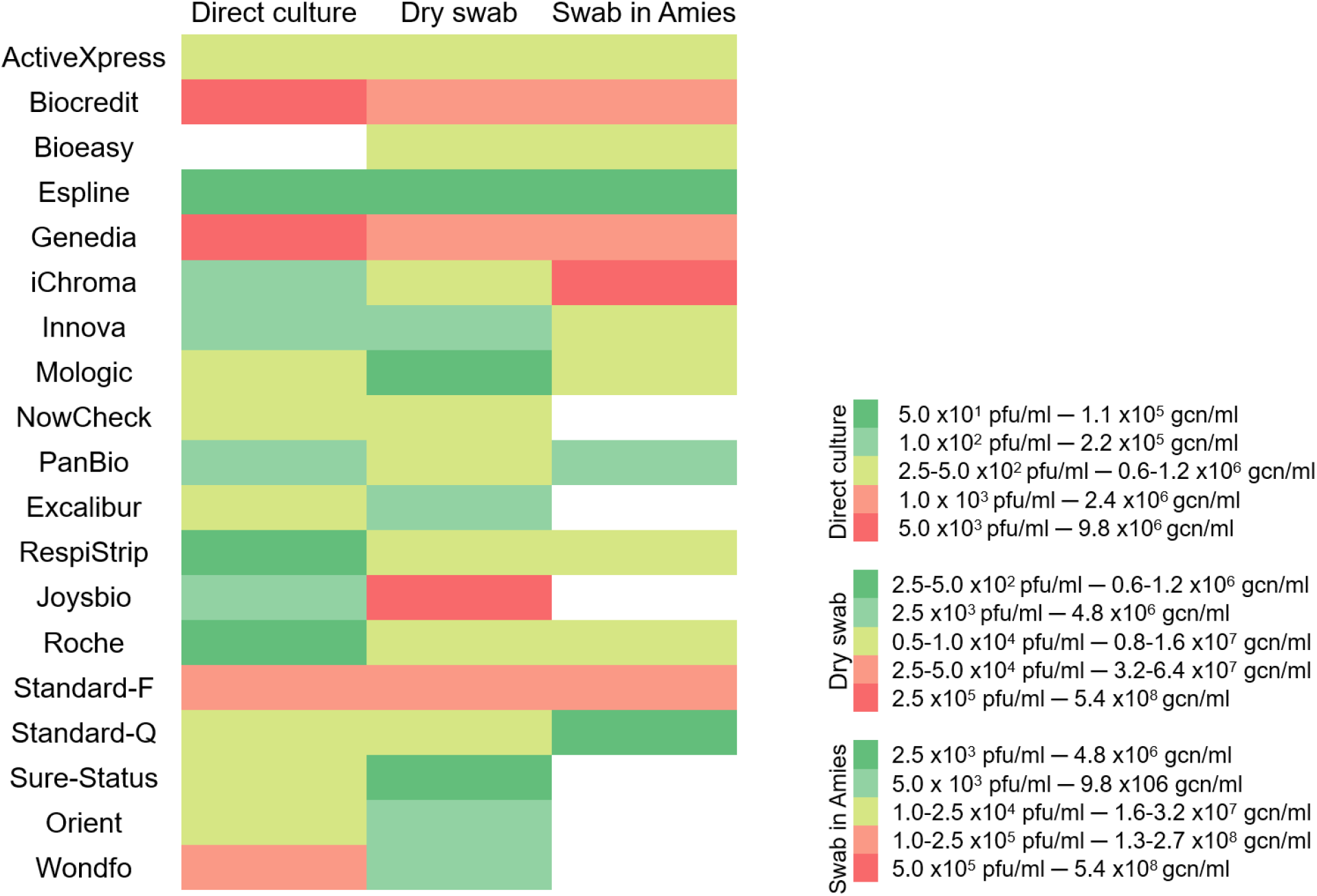
Heatmap of the LODs on all matrices. Ag-RDTs in direct culture matrix colored green fulfilled WHO criteria. Note: no colored cells for direct culture indicates no Ag-RDT was tested with that matrix, no colored cells in swab in Amies swab means interference with that matrix, hence LOD is not available.

### Volume of liquid recovered by swabs

We investigated whether the absorbance of the proprietary swabs provided with the Ag-RDT kits affected the LOD compared with direct culture supernatant i.e. if a less absorbent swab resulted in a poorer LOD in dry swab compared with the LOD obtained in direct culture supernatant for the same test. To test this hypothesis a Spearman’s correlation coefficient was performed comparing direct culture supernatant and dry swab matrices from the same Ag-RDT. The volume recovered by swabs per tests are shown in supplementary materials. Spearman’s correlation coefficient did not show any statistically significant correlation between the LOD and the volume recovered by swabs (P=0.421, ρ= −0.50), volume of extraction buffer (P=0.483, ρ= − 0.011) and dilution factor (calculated by considering the volume recovered by swabs and volume of extraction buffer) (P=0.460, ρ=-0.025).

### LOD one freeze-thaw cycle after 7 days at −80°C

Ag-RDTs are intended to be POC tests and thus the majority are recommended for use with freshly collected specimens. To validate test performance, use of stored material is much easier. Therefore, we performed this experiment to understand whether LOD is impacted following sample storage. The LOD of each of the tests using SARS-CoV-2 serial dilutions after 7 days storage at −80°C and one freeze-thaw cycle is shown in Table 2.

The LOD for 6 tests (ActiveXpress, Bioeasy, iChroma, NowCheck, RespiStrip and Wondfo) was equivalent using stored, frozen viral dilutions compared to fresh preparations. Three tests showed increased sensitivity (lower LOD by 2 to 5-fold) (Joysbio, PanBio and Standard-Q), and eleven showed poorer performance with a higher LOD of 2-fold (Biocredit, Espline, Roche and Sure-Status), 4-5 (Excalibur, Mologic and Orient), ten (Innova) and twenty (Genedia and Standard-F).

## Discussion

Here, we present the analytical performance of nineteen antigen rapid tests, which are currently on the market and in use in multiple countries. Analytical LODs are a useful proxy of clinical sensitivity, and the most standardized way to evaluate multiple antigen tests head-to-head, as each test requires a separate swab from patients. An approximate LOD of ≤ 5.0 × 10^2^ pfu/ml (≈ 1.0 × 10^6^ copies/ml) calculated using direct culture supernatant, has been proposed as the minimal analytical sensitivity by the WHO and the Department of Health and Social Care (DHSC, U.K.) ^25,26^. Fourteen of the nineteen marketed Ag-RDTs evaluated in this study fulfill this requirement (ActiveXpress, Bioeasy, Espline, Innova, Mologic, NowCheck, PanBio, Excalibur, RespiStrip, Joysbio, Roche, Standard-F, Standard-Q, Sure-Status and Orient).

Evaluation of the LOD using the kit-specific swabs immersed in the viral culture dilutions offers a more representative comparison to the level of sensitivity for clinical samples than using direct viral culture. Only four of the nineteen tests detected samples with concentrations ≤ 5.0 × 10^2^ pfu/ml (≈ 1.0 × 10^6^ gcn/ml) when using the dry swabs (Mologic, Espline, Roche, Sure-Status) and none of the tests met that LOD target when using swabs in Amies, like due to the dilution factor with the addition of 1 ml Amies buffer as well as potential chemical interactions between the media and the kit-specific buffers.

The more absorbent swabs will absorb more viral material, and so we investigated if the volume retained by the swab had any bearing on the LOD of each test when compared with the LOD achieved with direct culture supernatant. We took into account the volume recovered from the swab and the volume of proprietary buffer provided, but no correlation was found, this suggests that other factors may reduce the sensitivity when using swabs such as differences in the formulation of the proprietary buffers. A reduction in sensitivity using clinical samples may be observed compared to swabs in viral culture, as clinical samples are more viscous than culture media, potentially resulting in less viral material being absorbed onto the swab. The efficiency of the recovery is also likely to be increased by the centrifugation method used in our protocol.

We also evaluated the LOD and compatibility of the Ag-RDTs using a swab placed in Amies media, as these are routinely used to collect upper-respiratory samples in SARS-CoV-2 suspected individuals for diagnosis using RT-PCR^27,28^. If the same swab proves to be suitable for both RT-PCR and rapid antigen testing, one swab can be used for both tests as part of a serial algorithm. As well, the frozen leftover Amies media from RT-PCR testing could be used for future Ag-RDT evaluations. Either approach could simplify clinical and/or evaluation workflows. None of the Ag-RDT manufacturers specifically recommend the use of Amies media, and we demonstrated here that this ‘off-label’ sample preparation should be used with caution: six tests had false positive results (Espline, Excalibur, Joysbio, Sure-Status and Orient) and sensitivity was also reduced due to the additional volume of Amies.

The effect of storage at −80°C and one freeze-thaw cycle was evaluated, with eleven Ag-RDTs showing a loss of sensitivity by up to twenty-fold. A small decrease in sensitivity has been reported in in SARS-CoV-2 RT-PCR testing (<1 RT-PCR cycle threshold) after one and two freeze-thaw cycles ^29^ but there are no studies so far that have reported the effect of freeze-thaw on antigen detection. Results here highlight that the use of frozen material with Ag-RDTs should be performed with caution. The fact that three tests showed two-to-five-fold better sensitivity after an additional freeze thaw-cycle could not be explained in here, a further investigation is required with a larger sample size to rule out whether this phenomenon was within the margin of error of the experiment.

Three out of nineteen Ag-RDTs (Bioeasy, iChroma and Standard-F) rely on detection of a fluorescent signal using a reader. Though this may enable quantitative detection and potentially more consistent result interpretation, we did not find any improved sensitivity for this test format. Furthermore, Ag-RDTs that rely on a device may limit testing throughput if only one test can be read at a time. The reader also presents additional costs, as well as potential technical and maintenance issues which can be a barrier to implementation.

This analytical study has some limitations, as only a single isolate (REMRQ0001/Human/2020/Liverpool) was used to assess the LODs but our results are consistent with other recently-published analyses^30,31^. To the authors knowledge, all 19 tests evaluated here detect the nucleoprotein, presumably chosen for abundance and relative low mutation rate and therefore hypothesized to pick up all currently known variants^32,33^. Another limitation is that only one lot per kit was evaluated.

There is a growing number of studies suggesting that although antigen detection is less analytically sensitive than nucleic acid amplification techniques, it may strongly correlate with culturable virus, which may be a proxy for transmissibility. Hence Ag-RDTs could be informative for test, trace, isolate processes for the most infectious individuals^10–15^. Viral loads have been estimated to range from 10^8^ to 10^11^ gcn/ml in the most infectious patients^34–36^. The majority of Ag-RDTs evaluated here have an LOD predicted to successfully diagnose infected individuals with higher viral loads in this range across all matrices, except Joysbio that had an LOD of 5.4 × 10^8^ gcn/ml in dry swab. Further, Biocredit, iChroma, Standard-F and Genedia tests had LOD greater than 1.0 × 10^8^ gcn/ml when using swabs placed in Amies and In conclusion, the most sensitive tests with an LOD ≤ 5.0 × 10^2^ pfu/ml (≈ 1.2 ×10^6^ gcn/ml) on dry swabs and direct culture supernatant were Espline, Mologic, Sure-Status and Roche and the least sensitive on all matrices were Biocredit, iChroma, Standard-F and Genedia. The differences of LODs found here between tests and/or matrices ranged between 2-3 logs (i.e. 100-1,000 fold). Some tests showed impaired performance when using freeze-thaw material and/or Amies media. These findings highlight the importance of understanding assay specific performances and the need to select the appropriate sample matrix and the right test for each intended use, particularly for laboratories and evaluation programs that seek a rapid validation of Ag-RDT using frozen stored samples and ‘off-label’ specimen preparations. The LODs obtained in this comprehensive assessment of analytical sensitivity are consistent with rapidly emerging clinical performance data^9,11,37^ demonstrating the high clinical accuracy of Ag-RDTs for rapid detection of individuals with high viral loads, which can be very impactful for initiation of isolation and tracing measures.

## Methods

### Evaluated Ag-RDTs

Nineteen Ag-RDT based on lateral flow principle were evaluated in this study (Table 1): (1) ActivXpress+ COVID-19 Ag Complete Kit (Edinburgh Genetics Ltd), referred to as ActivXpress+, (2) Biocredit COVID-19 Ag (Rapidgen Inc.), referred to as Biocredit, (3) Bioeasy 2019-nCoV Ag (Shenzhen Bioeasy Biotechnology), referred to as Bioeasy, (4) Espline® SARS-CoV-2 (Fujirebio Diagnostics Inc.), referred to as Espline, (5) Genedia W COVID-19 Ag (Green Cross Medical Science), referred to as Genedia, (6) iChroma COVID-19 Ag Test (Boditech Medical Inc.), referred to as iChroma, (7) Innova SARS-CoV-2 Antigen Rapid (Innova Medical Group Ltd.), referred to as Innova, (8) Mologic COVID-19 Ag Test device (Mologic Ltd), referred to as Mologic, (9) NowCheck COVID-19 Ag test (Bionote Inc.), referred to as NowCheck, (10) Panbio™ COVID-19 Ag Rapid Test (Abbott Rapid Diagnostics), referred to as Panbio, (11) Rapid SARS-CoV-2 Antigen test card (Excalibur Healthcare Services), referred to as Excalibur, (12) Respi-Strip COVID-19 Ag (Coris Bioconcept), referred to as Respi-Strip, (13) SARS-CoV-2 Antigen Rapid Test Kit (Joysbio Biotechnology Ltd.), referred to as Joysbio, (14) SARS-CoV-2 Rapid Antigen Test (co-developed by SD Biosensor Inc and Roche Diagnostics, distributed by Roche Diagnostics), referred to as Roche, (15) Standard-F COVID-19 Ag FIA (SD Biosensor Inc), referred to as Standard-F, (16) Standard-Q COVID-19 (SD Biosensor Inc), referred to as Standard-Q, (17) Sure Status COVID-19 Antigen Card Test (Premier Medical Corporation), referred to as Sure-Status, (18) Coronavirus Ag Rapid Test (Zhejiang Orient Gene Biotech Ltd.), referred to as Orient, (19) Wondfo 2019-nCoV Antigen Test (Guangzhou Wondfo Biotech Co.), referred to as Wondfo. The selection of the Ag-RDT was based on expression of interest via the Infection Innovation Consortium (iiCON) and Foundation of New Diagnostics (FIND). Companies had no involvement in the design or reporting of the study.

**Table 1.** Characteristics of the Ag-RDT tested.

| Test name in this study | Assay | Manufacturer/Distributor | Country | Recommended sample | Principle | Format | Minutes to result |
| --- | --- | --- | --- | --- | --- | --- | --- |
| ActiveXpress | ActivXpress+ COVID-19 Ag Complete Kit | Edinburgh Genetics Ltd | UK | NP/OP swab | Colloidal gold | Cassette | 15-20 |
| Biocredit | Biocredit COVID-19 Ag | Rapidgen Inc. | Rep. Korea | NP swab/UTM/VTM | Colloidal gold | Cassette | 30 |
| Bioeasy | Bioeasy 2019-nCoV Ag | Shenzhen Bioeasy Biotechnology | China | NP swab | Fluorescence | Cassette | 10 |
| Espline | ESPLINE® SARS-CoV-2 | Fujirebio Diagnostics Inc. | Japan | NP swab | Colloidal gold | Cassette | 30 |
| Excalibur | Rapid SARS-CoV-2 Antigen test card | Boson Diagnostics/Excalibur Healthcare Services | UK | NP swab | Colloidal gold | Cassette | 15 |
| Genedia | GENEDIA W COVID-19 Ag | Green Cross Medical Science. | Rep. Korea | NP swab/Sputum | Colloidal gold | Cassette | 5-10 |
| iChroma | iChroma COVID-19 Ag Test | Boditech Medical Inc. | Rep. Korea | NP swab | Fluorescence | Cassette | 12 |
| Innova | Innova SARS-CoV-2 Antigen Rapid | Innova Medical Group Ltd. | UK | N/T swab | Colloidal gold | Cassette | 15 |
| Joysbio | SARS-CoV-2 Antigen Rapid Test Kit | Joysbio Biotechnology Ltd. | China | N swab | Colloidal gold | Cassette | 15-20 |
| Mologic | Mologic COVID-19 Ag Test device | Mologic Ltd. | UK | T/N/NT swab | Colloidal gold | Cassette | 10 |
| NowCheck | NowCheck COVID-19 Ag test | Bionote Inc./Mologic Ltd. | Rep. Korea | NP swab | Colloidal gold | Cassette | 15-30 |
| Orient | Coronavirus Ag Rapid Test | Zhejiang Orient Gene Biotech Ltd. | China | NP swab | Colloidal gold | Cassette | 10-15 |
| PanBio | Panbio™ COVID-19 Ag Rapid Test | Abbott Rapid Diagnostics | Rep. Korea | NP swab | Colloidal gold | Cassette | 15 |
| RespiStrip | Respi-Strip COVID-19 Ag | Coris Bioconcept | Belgium | NP wash/UTM/VTM | Colloidal gold | Dipstick | 15-30 |
| Roche | SARS-CoV-2 Rapid Antigen Test | SD Biosensor Inc./ Roche Diagnostics) | Switzerland | NP swab/VTM/UTM | Colloidal gold | Cassette | 15-30 |
| Standard-F | Standard F COVID-19 Ag FIA | SD Biosensor Inc. | Rep. Korea | NP swab | Fluorescence | Cassette | 15-30 |
| Standard-Q | Standard Q COVID-19 | SD Biosensor Inc. | Rep. Korea | NP swab | Colloidal gold | Cassette | 15 |
| Sure-Status | Sure-Status COVID-19 Antigen Card Test | Premier Medical Corporation | India | NP swab | Colloidal gold | Cassette | 15-20 |
| Wondfo | Wondfo 2019-nCoV Antigen Test | Guangzhou Wondfo Biotech Co. | China | NP/OP swab/VTM | Colloidal gold | Cassette | 10-15 |

Abbreviations: Rep.: Republic, OP: oropharyngeal, NP: nasopharyngeal, N: nasal, T: throat, NT: nasal-throat, UTM: universal transport media, VTM: viral transport media

**Table 2.** LOD after one week −80°C storage and one freeze-thawed cycle.

| **Test** | **LOD dry swab** | **LOD after -80°C storage** |
| --- | --- | --- |
| ActiveXpress | $1.0 \times 10^4$ | $1.0 \times 10^4$ |
| Biocredit | $5.0 \times 10^4$ | $1.0 \times 10^5$ |
| Bioeasy | $5.0 \times 10^3$ | $5.0 \times 10^3$ |
| Espline | $5.0 \times 10^2$ | $1.0 \times 10^3$ |
| Genedia | $2.5 \times 10^4$ | $5.0 \times 10^5$ |
| lChroma | $1.0 \times 10^4$ | $1.0 \times 10^4$ |
| Innova | $1.0 \times 10^3$ | $1.0 \times 10^4$ |
| Mologic | $2.5 \times 10^2$ | $1.0 \times 10^3$ |
| NowCheck | $5.0 \times 10^3$ | $5.0 \times 10^3$ |
| PanBio | $5.0 \times 10^3$ | $2.5 \times 10^3$ |
| Excalibur | $1.0 \times 10^3$ | $5.0 \times 10^3$ |
| RespiStrip | $5.0 \times 10^3$ | $5.0 \times 10^3$ |
| Joysbio | $2.5 \times 10^5$ | $1.0 \times 10^5$ |
| Roche | $5.0 \times 10^2$ | $1.0 \times 10^3$ |
| Standard-F | $2.5 \times 10^4$ | $5.0 \times 10^5$ |
| Standard-Q | $5.0 \times 10^3$ | $1.0 \times 10^3$ |
| Sure-Status | $5.0 \times 10^2$ | $1.0 \times 10^3$ |
| Orient | $2.5 \times 10^3$ | $1.0 \times 10^4$ |
| Wondfo | $2.5 \times 10^3$ | $2.5 \times 10^3$ |

### SARS-CoV-2 serial dilutions and quantification of copy numbers

The SARS-CoV-2 isolate REMRQ0001/Human/2020/Liverpool was propagated in Vero E6 cells (C1008; African green monkey kidney cells), maintained in DMEM with 2% fetal bovine serum (FBS) and 0.05 mg/ml gentamycin. Ten-fold serial dilutions of SARS-CoV-2 stock were made starting from 1.0 × 10^6^ pfu/ml to 1.0 × 10^2^ pfu/ml using culture media as a diluent (DMEM with 2% FBS % and 0.05 mg/ml gentamycin). Two-fold dilutions were made below the ten-fold LOD dilution to refine the LOD. For quantification, viral RNA was extracted using QIAmp Viral RNA mini kit (Qiagen, Germany) according to the manufacturer’s instructions. The genome copies/ml (gcn/ml) were calculated using the COVID-19 Genesig RT-qPCR kit (PrimerDesign, UK). RT-qPCR testing was carried out using the Rotor-Gene Q (Qiagen, Germany), with a ten-fold serial dilution of using quantified specific in vitro-transcribed RNA^38^. A total of five replicates were tested for each standard curve point and extracted RNA from each culture dilution was tested in triplicate, and the gcn/ml was calculated from the mean Ct value of these replicates.

### Preparation of SARS-CoV-2 sample matrices and LOD testing protocol

Three types of sample matrices were tested 1) direct viral culture supernatant, 2) spiked dry swabs and 3) spiked wet swabs in Amies media.

For the direct viral culture matrix, a specific volume of the serial dilutions was added directly to the extraction buffers at a 1:10 ratio except for Respi-Strip which was added at 1:1 ratio with the extraction buffer following the IFU.

For dry swab testing, the proprietary nasopharyngeal (NP) or nasal (N) swabs included in each individual kit was used except for Respi-Strip, which does not include swabs, and the recommended Eswab (Copan, Italy) was used instead. To prepare the dry swab matrix, the swab was soaked in 1 ml of the virus culture dilution series for 6-8 seconds, followed by immersion in the prescribed amount of proprietary reaction buffer solution.

For the preparation of spiked wet swabs, Eswab in Amies media (Copan, Italy) was used across all tests. The swab was first immersed in the serial viral dilutions for 6-8 seconds, then placed into the Amies media to mimic the sample collection stage. Ag-RDTs were evaluated by then immersing the same swab into the extraction buffer, except for test Respi-Strip where 100µl of the Amies was mixed at 1:1 with the extraction buffer following its IFU.

For all Ag-RDTs and matrices, the sample volumes applied, and procedures were performed as specified in the test specific IFUs.

The LOD was defined as the lowest dilution at which all three replicates were positive. Every dilution was tested in triplicate and non-spiked culture media and Amies were used as negative controls. Results were interpreted by two operators, each blinded to the result of the other. If a discrepant result was obtained, a third operator read any discrepant tests for a 2/3 result.

### Volume of liquid recovered by swabs

We investigated the effect of the absorbance of the proprietary swabs provided with the Ag-RDT kit in the LOD using dry swab, i.e. if a less absorbent swab resulted in a poorer LOD on dry swab compared with direct culture supernatant. To compare the effectiveness of each NP and N swab to recover sample, the amount of liquid absorbed by the swabs was measured. Five replicates of each swab brand were immersed in culture media for 6-8 seconds, then taped on the inside of a 50ml centrifuge tube. These were then centrifuged for 5 minutes at 1000g and the amount of liquid released was measured using a micropipette. The dilution factor of each test type was calculated taking into account the volume recovered by swab and volume of proprietary buffer in which the swab was immersed.

The degree of correlation of LOD by swabs with the volume recovered by swab type, volume of proprietary buffer and dilution factor were investigated by Spearman’s correlation coefficient rho (ρ). Statistical significance was set at P <0.05.

### LOD after 7 days at −80° C and one freeze-thaw cycle

After performing the LOD experiments, the viral culture dilutions were stored at –80 °C for 7 days and then the LOD experiments were performed again using the dry NP and N swabs. This would help to assess the use of stored clinical samples could be used to facilitate evaluation of Ag-RDTs.

## Data Availability

All data generated during this study is presented in an analyzed format in this manuscript. Raw datasets are available from the corresponding author on reasonable request.

## Data availability

All data generated during this study is presented in an analysed format in this manuscript. Raw datasets are available from the corresponding author on reasonable request.

## Authors’ contributions

The study and design of the study was conceived by AICA, JS and ERA. Laboratory testing was performed by AICA, KK, TE, CW, DW, KB, CT. Data analysis and interpretation were conducted by AICA. The initial manuscript was prepared by AICA. All authors edited and approved the final manuscript.

## Compelling Interest

The authors have no conflicts of interest to declare.

## References

1. Corman, V., Bleicker, T., Brünink, S. & Drosten, C. Diagnostic detection of 2019-nCoV by real-time RT-PCR. https://www.who.int/docs/default-source/coronaviruse/protocol-v2-1.pdf (2020).

2. Akst, J. RNA Extraction Kits for COVID-19 Tests Are in Short Supply in US. The Scientist https://www.the-scientist.com/news-opinion/rna-extraction-kits-for-covid-19-tests-are-in-short-supply-in-us-67250 (2020).

3. Kuznia, R., Curt, D. & Griffin, D. Severe shortages of swabs and other supplies hamper coronavirus testing - CNN. CNN US https://www.cnn.com/2020/03/18/us/coronovirus-testing-supply-shortages-invs/index.html.

4. Reagents hold up European COVID-19 tests. C&EN Glob. Enterp. (2020) doi:10.1021/cen-09813-buscon3.

5. Weekly statistics for NHS Test and Trace (England): 7 January to 13 January 2021. GOV.UK (2021).

6. Kretzschmar, M. E. et al. Impact of delays on effectiveness of contact tracing strategies for COVID-19: a modelling study. Lancet Public Heal. (2020) doi:10.1016/S2468-2667(20)30157-2.

7. World Health Organization (WHO). Antigen-detection in the diagnosis of SARS-CoV-2 infection using rapid immunoassays. Interim Guid. (2020).

8. Adams, E. R. et al. Rapid development of COVID-19 rapid diagnostics for low resource settings: accelerating delivery through transparency, responsiveness, and open collaboration. medRxiv (2020) doi:10.1101/2020.04.29.20082099.

9. Krüger, L. J. et al. Evaluation of the accuracy, ease of use and limit of detection of novel, rapid, antigen-detecting point-of-care diagnostics for SARS-CoV-2. medRxiv (2020).

10. Weiss, G. & Bellmann-Weiler, R. Rapid antigen testing and non-infectious shedding of SARS-Cov2. Infection (2021) doi:https://doi.org/10.1007/s15010-020-01570-w.

11. Iglὁi, Z. et al. Clinical evaluation of the Roche/SD Biosensor rapid antigen test with symptomatic, non-hospitalized patients in a municipal health service drive-through testing site. medRxiv (2020).

12. Mina, M. J., Parker, R. & Larremore, D. B. Rethinking Covid-19 Test Sensitivity — A Strategy for Containment. N. Engl. J. Med. (2020) doi:10.1056/nejmp2025631.

13. Options for the use of rapid antigen tests for COVID-19in the EU/EEAand the UK. Eur. Cent. Dis. Prev. Control. (2020).

14. Pekosz, A. et al. Antigen-based testing but not real-time PCR correlates with SARS-CoV-2 virus culture. medRxiv (2020).

15. Singanayagam, A. et al. Duration of infectiousness and correlation with RT-PCR cycle threshold values in cases of COVID-19, England, January to May 2020. Eurosurveillance (2020) doi:10.2807/1560-7917.ES.2020.25.32.2001483.

16. Abduljalil, J. M. Laboratory diagnosis of SARS-CoV-2: available approaches and limitations. New Microbes and New Infections (2020) doi:10.1016/j.nmni.2020.100713.

17. Bonačić Marinovic, A. A. et al. Speed versus coverage trade off in targeted interventions during an outbreak. Epidemics (2014) doi:10.1016/j.epidem.2014.05.003.

18. Mahase, E. Covid-19: Mass testing in Slovakia may have helped cut infections. BMJ (2020) doi:10.1136/bmj.m4761.

19. Gill, M. & Gray, M. Mass testing for covid-19 in the UK. The BMJ (2020) doi:10.1136/bmj.m4436.

20. Wise, J. Covid-19: Government ramps up”Moonshot”mass testing. BMJ 371, (2020).

21. Mak, G. C. et al. Evaluation of rapid antigen test for detection of SARS-CoV-2 virus. J. Clin. Virol. (2020) doi:10.1016/j.jcv.2020.104500.

22. Nagura-Ikeda, M. et al. Clinical evaluation of self-collected saliva by quantitative reverse transcription-PCR (RT-qPCR), Direct RT-qPCR, reverse transcription-loop-mediated isothermal amplification, and a rapid antigen test to diagnose COVID-19. J. Clin. Microbiol. (2020) doi:10.1128/JCM.01438-20.

23. Scohy, A. et al. Low performance of rapid antigen detection test as frontline testing for COVID-19 diagnosis. J. Clin. Virol. (2020) doi:10.1016/j.jcv.2020.104455.

24. Weitzel, T. et al. Head-to-head comparison of four antigen-based rapid detection tests for the diagnosis of SARS-CoV-2 in respiratory samples. bioRxiv (2020) doi:10.1101/2020.05.27.119255.

25. gov.uk. Protocol for evaluation of rapid diagnostic assays for specific SARS-CoV- 2 antigens (lateral flow devices).

26. (WHO), W. H. O. & R&D Blue Print, W. H. (HQ). Target product profiles for priority diagnostics to support response to the COVID-19 pandemic v.1.0. (2020).

27. Center of Disease Control and Prevention. Interim Guidelines for Collecting, Handling, and Testing Clinical Specimens for COVID-19. May 22 (2020).

28. Federman, D. G. et al. SARS-CoV-2 detection in setting of viral swabs scarcity: Are mrsa swabs and viral swabs equivalent? PLoS One (2020) doi:10.1371/journal.pone.0237127.

29. Li, L. et al. Influence of Storage Conditions on SARS-CoV-2 Nucleic Acid Detection in Throat Swabs. J. Infect. Dis. (2020) doi:10.1093/infdis/jiaa272.

30. Corman, V. M. et al. Comparison of seven commercial SARS-CoV-2 rapid Point-of-Care Antigen tests. medRxiv (2020) doi:10.1101/2020.11.12.20230292.

31. Jääskeläinen, A. et al. Evaluation of three rapid lateral flow antigen detection tests for the diagnosis of SARS-CoV-2 infection. medRxiv (2021).

32. Bar-On, Y. M., Flamholz, A., Phillips, R. & Milo, R. Sars-cov-2 (Covid-19) by the numbers. Elife (2020) doi:10.7554/eLife.57309.

33. Dutta, N. K., Mazumdar, K. & Gordy, J. T. The Nucleocapsid Protein of SARS– CoV-2: a Target for Vaccine Development. J. Virol. (2020) doi:10.1128/jvi.00647-20.

34. Kawasuji, H. et al. Transmissibility of COVID-19 depends on the viral load around onset in adult and symptomatic patients. PLoS One (2020) doi:10.1371/journal.pone.0243597.

35. Zou, L. et al. SARS-CoV-2 Viral Load in Upper Respiratory Specimens of Infected Patients. N. Engl. J. Med. (2020) doi:10.1056/NEJMc2001737.

36. Walsh, K. A. et al. SARS-CoV-2 detection, viral load and infectivity over the course of an infection. Journal of Infection (2020) doi:10.1016/j.jinf.2020.06.067.

37. van der Moeren, N. et al. Performance evaluation of a SARS-COV-2 rapid antigentest: Test performance in the community in the netherlands. medRxiv (2020) doi:10.1101/2020.10.19.20215202.

38. Byrne, R. L. et al. Saliva offers a sensitive, specific and non-invasive alternative to upper respiratory swabs for SARS-CoV-2 diagnosis. medRxiv (2020) doi:10.1101/2020.07.09.20149534.

